# Brain-rejuvenating factor TIMP2 is associated with brain health and neuroprotective lifestyle in aged subjects

**DOI:** 10.64898/2025.12.24.25342952

**Authors:** Emily W. Paolillo, Samuele F. Petridis, Ana Catarina Ferreira, Hanxiao Liu, Jeffrey D. Zhu, Rowan Saloner, Anna M. Vandebunte, Claire J. Cadwallader, Coty Chen, Joel H. Kramer, Kyan Younes, Maya V. Yutsis, Victor W. Henderson, David A. Bennett, Joseph M. Castellano, Kaitlin B. Casaletto

## Abstract

**Background:** Tissue inhibitor of metalloproteinases 2 (TIMP2) has been shown to revitalize aspects of synaptic plasticity and hippocampus-dependent cognition in aged mice. We examined TIMP2’s relevance in human brain aging and tested whether TIMP2 may be modifiable via known pro-plasticity behaviors in humans and mice.

**Methods:** Plasma TIMP2 levels were quantified via SomaScan in three independent human cohorts, including two cross-sectional (UCSF: n=83; Stanford: n=31) and one with longitudinal measurement (ROSMAP: n=213). We examined associations of plasma TIMP2 with cognitive performance, brain volumes, and engagement in neuroprotective lifestyle behaviors, including objectively measured physical activity (UCSF) and a multi-domain lifestyle composite (ROSMAP). We used mouse models to directly test effects of environmental enrichment on (1) plasma TIMP2 levels and (2) hippocampal neurogenesis in wildtype versus TIMP2 knockout mice.

**Results:** Higher plasma TIMP2 associated with better global cognition and larger brain volumes across human cohorts. Longitudinal decreases in plasma TIMP2 associated with steeper cognitive decline. Physical activity positively associated with plasma TIMP2 cross-sectionally, and longitudinal changes in multi-domain lifestyle factors positively associated with change in TIMP2 levels over time in humans. Mice exposed to an enriched environment for 3 weeks exhibited elevated plasma TIMP2 levels. While wildtype mice exposed to enrichment exhibited elevated adult hippocampal neurogenesis, this effect was lost in mice in which TIMP2 had been deleted.

**Conclusions:** TIMP2 demonstrates clinical relevance for brain aging in humans and may represent a mechanism through which lifestyle behaviors confer neuroprotection. Further examination of TIMP2 as a potential therapeutic target for prevention of cognitive decline is warranted.

## INTRODUCTION

Aging is accompanied by decline in cognitive function and elevated risk for neurodegenerative disease and other age-related neurological conditions (1). As the population of older adults continues to increase, there is an urgent need to identify therapeutics for maintaining brain health across the lifespan. Studies examining the young systemic compartment have highlighted specific proteins that could serve as therapeutic targets for supporting brain health into older adulthood (2, 3). One such candidate is tissue inhibitor of metalloproteinases-2 (TIMP2), a protein that regulates extracellular matrix (ECM) remodeling, inflammatory responses, angiogenesis, and tissue homeostasis (4) that has been previously identified as a therapeutic target and prognostic biomarker in cancer research (5).

TIMP2 is widely expressed in the body, crosses into brain from the blood (6), and has more recently been shown to support synaptic plasticity, adult hippocampal neurogenesis, and cognitive function in rodents (7). Previous work has shown that systemic administration of TIMP2 in aged mice enhances hippocampal function and improves memory performance, suggesting a rejuvenating effect on the aging brain (6). Models in which TIMP2 is genetically deleted demonstrate that neuronal TIMP2 directly regulates adult neurogenesis, dendritic spine plasticity, and hippocampus-dependent memory function (7). Together, these findings suggest that TIMP2 may have therapeutic potential for maintenance of brain health and dementia prevention; however, translational evidence for the beneficial effects of circulating levels of TIMP2 in humans is limited.

In parallel, modifiable lifestyle behaviors such as physical activity, cognitive engagement, and social interaction are well-established resilience factors against cognitive decline and dementia (8, 9). However, the molecular mechanisms underlying the neuroprotective effects of these lifestyle behaviors are not well understood. Given TIMP2’s role in angiogenesis and synaptic plasticity, processes that underlie the cognitive benefits of healthy lifestyle behaviors (10–12), we examined whether TIMP2 may be modifiable and underlies the neuroprotective effects of healthy lifestyle activities.

We leveraged three independent human cohorts and animal models to examine: 1) the translational relevance of plasma TIMP2 levels for human brain health, 2) whether plasma TIMP2 associates with engagement in neuroprotective lifestyle behaviors in humans, and 3) the relationship between lifestyle enrichment on TIMP2 levels and synaptic plasticity in mouse models (**Figure 1**). This study aims to bridge preclinical findings on TIMP2 with deeply-phenotyped human cohorts to uncover potential mechanisms through which lifestyle shapes neurobiological aging.

**Fig. 1.**
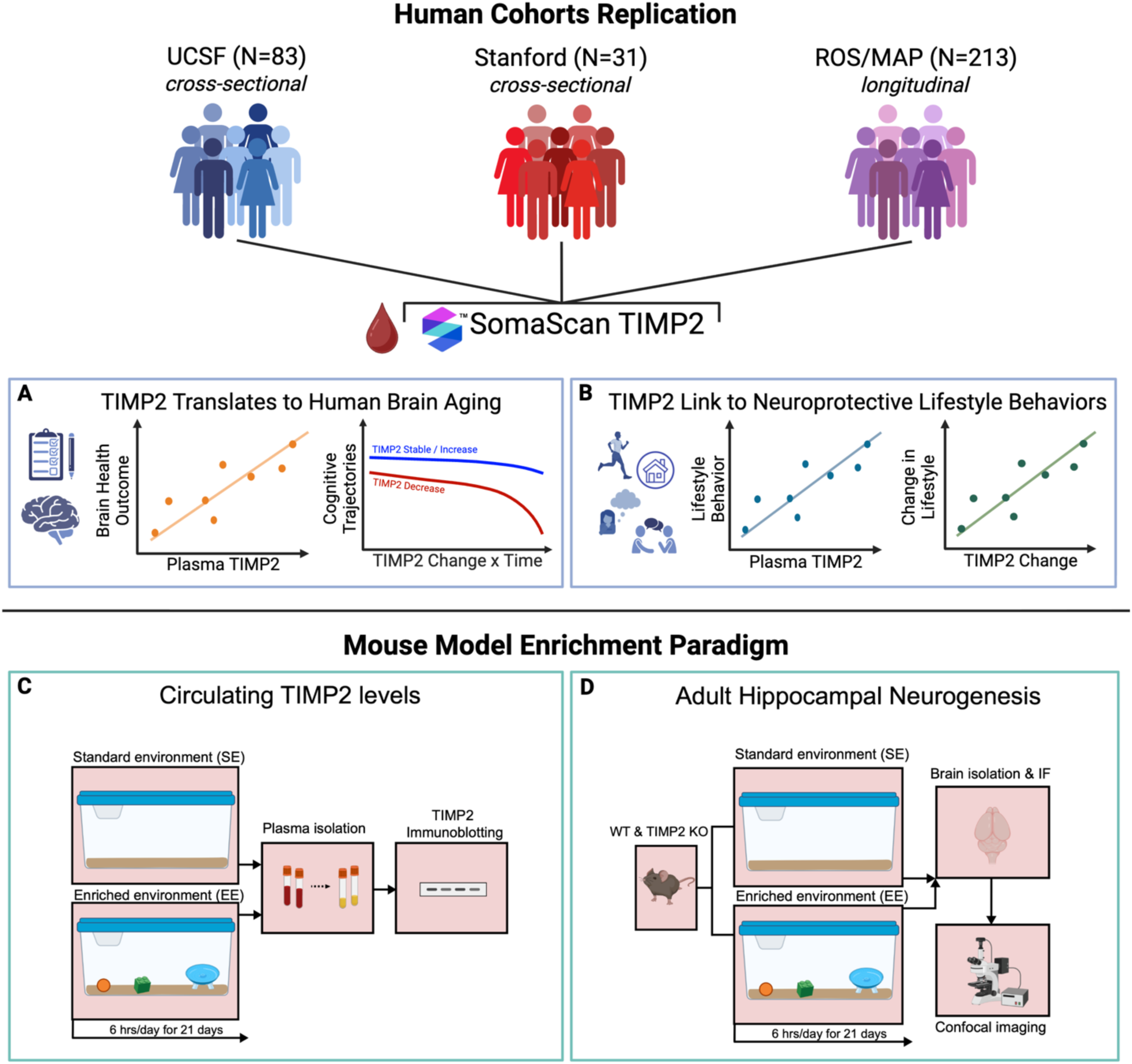
Study overview. Plasma TIMP2 was analyzed by SomaLogic in three independent human cohorts, with goals to **(A)** examine translational relevance of TIMP2 in human brain aging and **(B)** link TIMP2 to known brain rejuvenation behaviors. Given observed associations in humans, we examined causal relationships between lifestyle enrichment and TIMP2 in mouse models. We tested whether **(C)** exposure to an enriched environment influenced plasma TIMP2 and **(D)** the pro-neurogenic effects of environmental enrichment on the adult hippocampus were dependent on the presence of TIMP2.

## METHODS AND MATERIALS

### Human Participants

#### Cohort 1 (UCSF)

Participants included 83 community-dwelling older adults enrolled in the Brain Aging Network for Cognitive Health (BrANCH) and Alzheimer’s Disease Research Center (ADRC) studies at the UCSF Memory and Aging Center (MAC). Participants underwent comprehensive clinical evaluations including clinical interview, neurologic exam, neuropsychological testing, neuroimaging (MRI and amyloid PET), informant-based functional assessment (Clinical Dementia Rating), and blood draw. All participants also completed 30 days of Fitbit^TM^ Flex 2 monitoring. Per multidisciplinary consensus case conference, participants were categorized as either: 1) clinically normal; or 2) mild cognitive impairment (MCI) due to Alzheimer’s disease including amyloid PET positivity. All study procedures were approved by the UCSF Committee on Human Research and all participants provided written informed consent.

#### Cohort 2 (Stanford University)

31 participants from Stanford ADRC were selected for inclusion based on clinical diagnosis (clinically normal or MCI) and data availability for plasma TIMP2, AD biomarkers (CSF Aβ42/40), cognitive testing, brain MRI, and demographics. All study procedures were approved by the Stanford Committee on Human Research and all participants provided written informed consent.

#### Cohort 3 (Rush University)

213 participants from the Rush Alzheimer’s Disease Center (Religious Orders Study and Memory and Aging Project [ROSMAP]) were selected for inclusion based on data availability for plasma TIMP2 at *two timepoints* and longitudinal cognitive testing. ROSMAP participants also had longitudinal data on self-reported neuroprotective lifestyle factors (i.e., physical activity, social activity, cognitive activities, and life space) collected at each study visit and autopsy-based neuropathological data.

Together, these three clinical cohorts represent geographically distinct groups of both community-dwelling (UCSF, ROSMAP) and clinical (Stanford) cohorts with both cross-sectional (UCSF, Stanford) and longitudinal (ROSMAP) data (**Table 1**).

**Table 1.** Participant characteristics in each cohort. Characteristics in the ROSMAP cohort represent those at their baseline SomaLogic visit.

|  | UCSF<br>(N=83) | Stanford<br>(N=31) | ROSMAP<br>(N=213) |
| --- | --- | --- | --- |
| *Demographics* |  |  |  |
| Age | 75.7 (6.6) | 71.3 (6.5) | 82.6 (5.8) |
| Sex (women) | 48 (58%) | 12 (39%) | 155 (73%) |
| Race/Ethnicity |  |  |  |
| Non-Hispanic White | 70 (84%) | 27 (87%) | 206 (97%) |
| Asian | 9 (10%) | 2 (6%) | 0 (0%) |
| Black, Latinx, Multiracial, or Other | 4 (5%) | 2 (6%) | 7 (3%) |
| Education (years) | 17.3 (2.4) | 17.6 (2.0) | 14.4 (2.8) |
| *Clinical Characteristics* |  |  |  |
| Clinical Group |  |  |  |
| Cognitively normal | 65 (78%) | 26 (84%) | 213 (100%) |
| Mild cognitive impairment | 18 (22%) | 5 (16%) | 0 (0%) |
| Aβ-PET Status (positive) | 40 (48%) | - | - |
| Average Daily Step Count | 7319.9 (3271.8) | - | - |

Note. Values are mean (SD) or n (%)

### Plasma TIMP2

Plasma was collected per each study protocol and stored at −80°C until being packed with dry ice and sent to SomaLogic (SomaLogic, Boulder, CO) for proteomics analysis. Plasma proteins, including TIMP2, were measured on the SomaScan v4.1 platform in all cohorts via separate runs. SomaScan uses a volume of 65 microliters of plasma to create SOMAmer – protein reactions in 96-well plates. Tagged SOMAmer-protein complexes were captured in a bead-based assay, and levels of SOMAmer bound to sample were quantified through a fluorescent signal in DNA hybridization microarrays. The reaction signal was detected digitally and expressed as aggregated Agilent relative fluorescent units (RFU), which were normalized to scale and subsequently log2-transformed. In the ROSMAP cohort, change in TIMP2 levels was quantified as TIMP2 at baseline subtracted from TIMP2 at follow up. To test robustness across measurement techniques, we also assayed a subset of N=40 UCSF participants for plasma TIMP2 levels via enzyme-linked immunosorbent assay (ELISA; R&D Systems, Cat#DTM200, Minneapolis, MN, USA).

### Cognitive Functioning

UCSF participants completed a comprehensive neuropsychological battery, covering episodic memory, executive functioning, and language (13). Sample based z-scores were calculated for individual tests and averaged within each domain to create composite z-scores. Memory, executive functioning, and language z-scores were then averaged to create a global cognitive z-score. Stanford ADRC participants completed measures of episodic memory, executive functioning, and language (14). Similarly, sample-based z-scores were calculated for individual tests, averaged within each domain to create domain composite z-scores, then domain z-scores were averaged to create a global cognitive z-score. ROSMAP participants (15) completed measures of episodic memory, semantic processing, working memory, processing speed, and executive functioning longitudinally (median [IQR] of 5 [2-8] visits over a median [IQR] of 4 [1-7] years of follow-up). A global cognitive composite score was derived from a battery of 19 cognitive tests administered to participants each year (16). Raw scores from the 19 tasks were converted to z-scores, averaged within domains to produce domain z-scores, then averaged across domains to produce a global cognitive z-score, as previously described (16).

### Brain MRI and Amyloid PET

UCSF participants completed brain MRI using a Siemens Prisma 3T scanner. Acquisition and processing details have been published previously (15). Regions of interest (ROIs) from the Desikan atlas (17) were extracted and composite ROIs were computed for total gray matter volume. UCSF participants also completed Aβ-PET imaging with 18F-florbetapir (injected dose:≈10 mCi). Acquisition and processing details have been published previously (18). Each scan was identified as positive (reflecting elevated global cortical Aβ deposition) or negative (reflecting minimal cortical tracer uptake) by consensus visual read from a panel of three neurologists.

### Lifestyle Enrichment

To capture enrichment behaviors in humans, UCSF participants completed 30 days of Fitbit^TM^ Flex 2 monitoring within 1 year of their study visit. Participants were asked to wear the Fitbit continuously on their non-dominant wrist during all waking hours. All Fitbit activity feedback was disabled. Total daily step counts were collected. Days with fewer than 100 steps were excluded to remove noise from days when participants likely did not wear the device, per previously published procedures (19, 20). Total step counts from all remaining valid days were averaged for each participant as a measure of average daily activity levels. ROSMAP participants self-reported levels of *physical activity* (5-item hours of physical activity engagement in late life), *social activity* (6-item survey of common activities involving social interactions), *cognitive activity* (average frequency across seven cognitively stimulating activities in the past year), and *life space* (a proxy of environmental enrichment reflecting frequency of spatial movement in six increasingly distant environments, e.g., bedroom, neighborhood, out of town) via standardized measures at every visit. These measures have been extensively associated with cognitive aging and dementia risk in previous works (15, 21–26). A composite lifestyle enrichment score was derived by averaging sample-based z-scores across these four lifestyle measures with higher values indicating greater enrichment.

### Statistical Analysis of Human Data

Statistical analyses were performed using R, version 4. Pearson correlations or independent t tests examined relationships between plasma TIMP2 protein levels and demographics for continuous or dichotomous variables, respectively. Pearson correlation examined concordance between TIMP2 levels quantified across different platforms (i.e., SomaLogic vs. ELISA).

Multiple linear regression examined cross-sectional relationships of plasma TIMP2 protein levels with global cognitive z-scores and total gray matter volumes in the UCSF and Stanford cohorts, separately. Analyses covaried for age and Alzheimer’s disease pathology (amyloid PET status in UCSF; CSF Aβ42/40 ratio in Stanford); models predicting cognition additionally covaried for sex and years of education; models evaluating brain volumes additionally covaried for total intracranial volume. In ROSMAP, longitudinal data allowed us to evaluate how changes in plasma TIMP2 associated with cognitive trajectories (from first TIMP2 measurement to death); to do so, linear mixed effects regression examined longitudinal global cognitive functioning as a function of TIMP2 change (TIMP2 change x linear time [years before death]; TIMP2 change x quadratic time [years before death^2^]), covarying for baseline TIMP2 levels, age, sex, education, global AD burden at autopsy, total years of follow up (from first TIMP2 measurement to death), and each of their interactions with time. Person-specific random intercepts and a random effect of time were modeled.

Next, we tested how plasma TIMP2 levels associate with modifiable neuroprotective behaviors. In the UCSF cohort, multiple linear regression examined the relationship between plasma TIMP2 and physical activity. In ROSMAP, we examined the relationship between change in TIMP2 levels and change in overall lifestyle enrichment (baseline lifestyle composite score subtracted from lifestyle composite score at the TIMP2 follow up visit). Analyses covaried for age and AD pathology.

### Animal Experiments

Animal procedures were performed in accordance with the National Institutes of Health Guide for Care and Use of Laboratory Animals and the Icahn School of Medicine at Mount Sinai Institutional Animal Care and Use Committee. Mice were maintained on a C57Bl/6 background and TIMP2 knockout (KO) (Jackson Laboratory) and WT littermates were used at 2-3 months of age. Mice were maintained on a 12 h light/dark cycle at constant temperature (23°C) with ad libitum access to food and water.

### Cage Enrichment Paradigm

Mice were housed in a “home cage” from which mice were added to either an “enrichment cage” or “standard cage” during the exposure period. Enrichment cages contained an exercise saucer, a T-shaped tunnel that was hemisected to minimize mouse inactivity and/or sleep within the complete tunnel, colored block, variable toy balls, and a water bowl. Toy balls were exchanged daily to promote enrichment. Cages were given 8 randomly scattered food pellets before each exposure period. Enrichment cages were given variable arrangements that were alternated to maintain novelty for objects. Standard cages lacked enrichment but were identical in all other aspects.

Animals were randomly assigned to enrichment environment (EE) or standard environment (SE) control groups and subsequently exposed to these conditions for 6 hours per day for 21 days, adapted from previously established paradigms (27–29), starting at 12PM and ending at 6PM, corresponding to Zeitgeber time (ZT) ZT6 - ZT12. Cages were checked daily to confirm engagement with enrichment apparati, including the exercise wheel, and to ensure access was consistent for all mice. Heterozygous mice not used for study were retained as cagemates to maintain social enrichment. Following exposure, mice were returned to their respective home cages. Toys were cleaned daily with 70% ethanol and replaced according to the next unique cage orientation. Orientations were alternated according to a cycle throughout the exposure period. Mice were sacrificed 24 hours after the final exposure period, and brain tissue or blood was collected for respective downstream processing.

### Brain Immunohistochemistry

For sacrifice prior to immunohistochemistry experiments, mice were anesthetized with a cocktail of ketamine (90 mg/kg) and xylazine (10 mg/kg) and transcardially perfused with ice-cold 0.9% saline. Brains were post-fixed in 4% paraformaldehyde for 48 hours and preserved in 30% sucrose before sectioning at 40 µm on a freezing-sliding microtome (SM2010R, Leica). Sections were incubated overnight with primary antibody to stain immature neuroblasts (Doublecortin, 1:200, Cell Signaling, cat. #4604S) followed by respective fluorescent secondary antibody. Image processing was performed with LSM 780 confocal microscope (Zeiss) using 40×/1.4 Oil DIC objective. Four equally spaced sections per mouse were used to count the total number of DCX-positive cells within the subgranular zone of the dentate gyrus using stereological principles, as previously described (7, 30). All counts were performed in a blinded fashion using FIJI software.

### Plasma Immunoblotting for Mouse TIMP2

Blood was collected 24 hours following completion of the paradigm from EE- and SE-exposed mice by cardiac puncture (0.2-0.3 mL per mouse) and centrifuged at 4°C for 10 minutes to isolate EDTA-plasma. Plasma samples were stored at −80°C until use. For immunoblotting, equivalent volumes of plasma (2.5 μL) samples were loaded in a randomized fashion on 4-12% NuPAGE Bis-Tris gels (Invitrogen) for SDS-PAGE. Following electrophoresis, gels were transferred to nitrocellulose membranes, and blots were probed with anti-TIMP2 antibody (1:500, Cell Signaling, cat. #5738S) and developed as previously described (6). Plasma TIMP2 band intensities were quantified using FIJI in a blinded fashion. Ponceau S staining was used to assess confirm loading.

### Statistical Analysis of Mouse Data

Statistical analyses were performed using GraphPad Prism version 9.0 software (GraphPad Software) using tests described in figure legends. All experiments were performed in a blinded fashion, and randomization was performed where applicable. Some schematic illustrations were adapted from Biorender tools.

## RESULTS

### Plasma TIMP2 Levels Do Not Vary by Education, Amyloid status, or Other Demographic Factors in Older Adults

To begin to examine how TIMP2 levels vary in the plasma of older humans, we examined TIMP2 SomaLogic measurements from UCSF MAC (n=83; age range=56-93), Stanford University ADRC (n=31; age range=62-87), and Religious Orders Study (ROS) or Memory and Aging Project (MAP) at Rush University Medical Center (n=213; baseline ROSMAP age range=58-100; **Fig. 1**) studies. Plasma TIMP2 in UCSF subjects demonstrated weak, non-significant associations with age (r=0.11, 95%CI=-0.10 to 0.32, p=0.304), education (r=0.07, 95%CI=-0.15 to 0.28, p=0.561), and amyloid PET positivity (mean difference=0.04, t_76_=0.91, p=0.366). Males showed marginally higher TIMP2 levels (M=15.34 Log_2_ RFU, SD=0.17) than females (M=15.26 Log_2_ RFU, SD=0.21; t_81_=1.94, p=0.056). These demographic patterns were generally consistent across both Stanford and ROSMAP replication cohorts, such that plasma TIMP2 showed weak associations with age (Stanford: *r*=0.22, *p*=0.228; baseline ROSMAP; *r*=0.08, *p*=0.233), education (Stanford: *r*=0.30, *p*=0.106; baseline ROSMAP: *r*=0.03, *p*=0.676), and biomarkers for brain amyloid (Stanford [plasma Aβ42/40 ratio]: *r*=0.09, *p*=0.616; baseline ROSMAP [global AD pathology burden at autopsy]: *r*=0.09, *p*=0.216). In the Stanford cohort, males had higher plasma TIMP2 levels (M=15.60 RVU, SD=0.19) than females (M=15.48 RVU, SD=0.09; t_28_=2.35, p=0.026), while sex differences did not reach statistical significance in ROSMAP (males: M=15.36 RVU, SD=0.23; females: M=15.35 RVU, SD=0.24; *p*=0.722). In ROSMAP, change in TIMP2 levels associated with years between TIMP2 measurements (*r*=0.15, *p*=0.024), but did not significantly relate to baseline demographic factors (*p*s>0.05) nor global AD pathology burden at autopsy (*r*=-0.05, *p*=0.454).

We next examined TIMP2 measurements using an orthogonal platform. TIMP2 levels exhibited good cross-platform concordance in the subset of 40 UCSF participants who had plasma analyzed for TIMP2 across both SomaScan and ELISA (*r*=0.61, 95%CI=0.37 to 0.77, p<0.001; **Suppl. Fig. 1**), thereby validating aptamer-based measurement of plasma TIMP2.

### Plasma TIMP2 Levels Associate with Cognitive and Brain Outcomes in Humans

We next evaluated whether neuroprotective effects of plasma TIMP2 in aged animal models are recapitulated in older human adults. Plasma TIMP2 levels associated with better global cognitive functioning in both the UCSF (*Std. β*=0.232, *95%CI*=0.010 to 0.454, *p*=0.041; **Fig. 2A**) and Stanford cohorts (*Std. β*=0.382, *95%CI*=0.053 to 0.711, *p*=0.025; **Fig. 2B**). Similarly, higher plasma TIMP2 levels associated with larger gray matter volumes in UCSF (*Std. β*=0.174, *95%CI*=0.003 to 0.344, *p*=0.046; **Fig. 2C**) and Stanford cohorts (*Std. β*=0.200, *95%CI*=0.001 to 0.400, *p*=0.048; **Fig. 2D**). We next asked whether intra-individual changes in plasma TIMP2 were associated with changes in cognition over time in a longitudinal ROSMAP cohort. Changes in plasma TIMP2 were normally distributed (**Fig. 3A-B**). Decreases in plasma TIMP2 associated with steeper global cognitive declines (TIMP2 x time: *Std. β*=0.111, *95%CI*=0.001 to 0.220, *p*=0.047; TIMP2 x time^2^: *Std. β*=0.091, *95%CI*=-0.010 to 0.192, *p*=0.079; **Fig. 3C**), which was primarily driven by changes in episodic memory performance (TIMP2 x time: *Std. β*=0.203, *95%CI*=0.082 to 0.323, *p*=0.001; TIMP2 x time^2^: *Std. β*=0.174, *95%CI*=0.058 to 0.291, *p*=0.003; **Fig. 3D**; see **Supplementary Tables 1-4** for full model results from each cohort). Together, data from three distinct cohorts converge to support translational relevance of plasma TIMP2 for cognitive aging in humans.

**Fig. 2.**
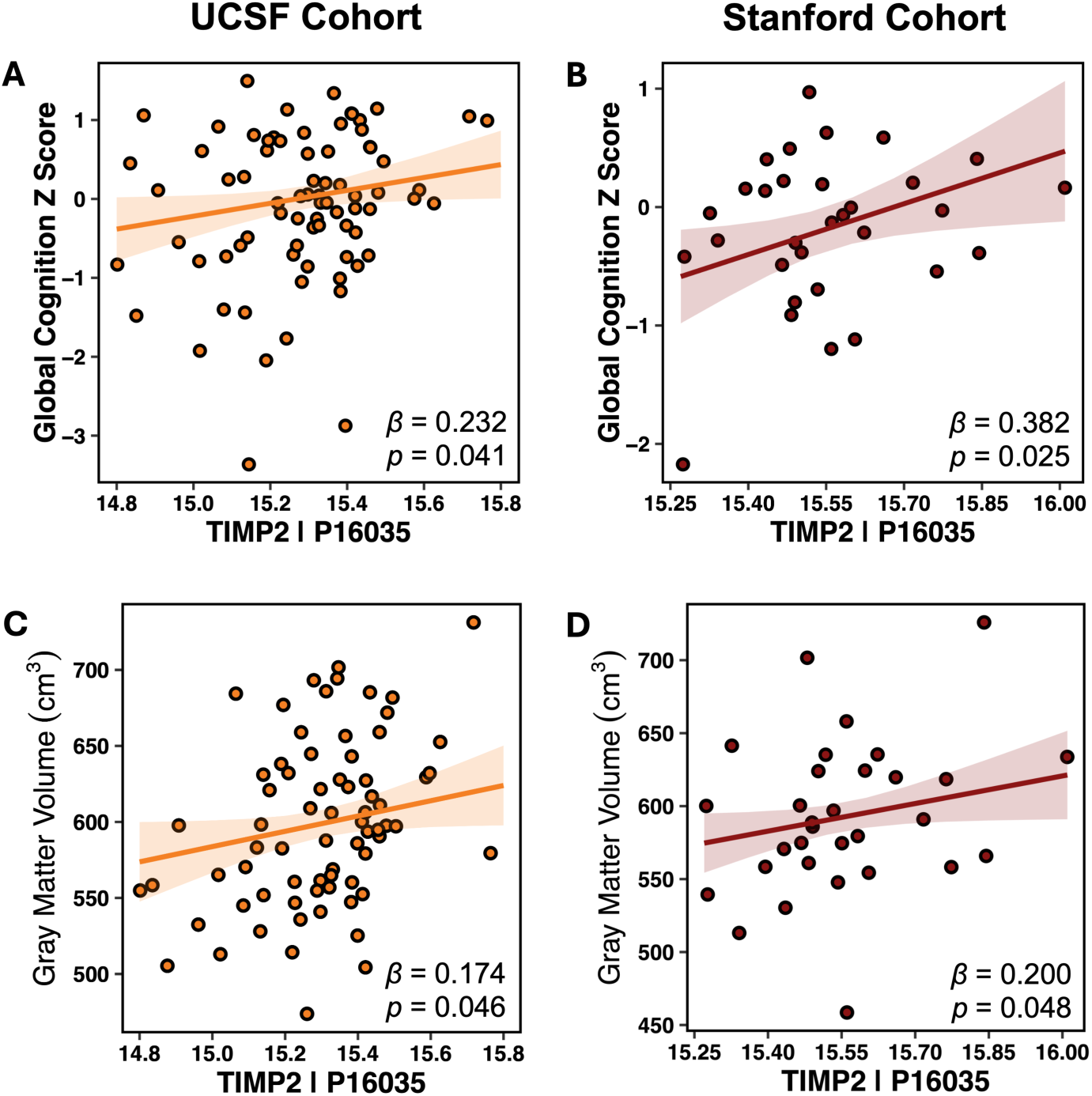
Higher levels of TIMP2 relate to better cognitive and brain aging in humans. Plasma TIMP2 is positively associated with global cognition **(A,B)** and total gray matter volumes **(C,D)** in both UCSF and Stanford cohorts.

**Fig. 3.**
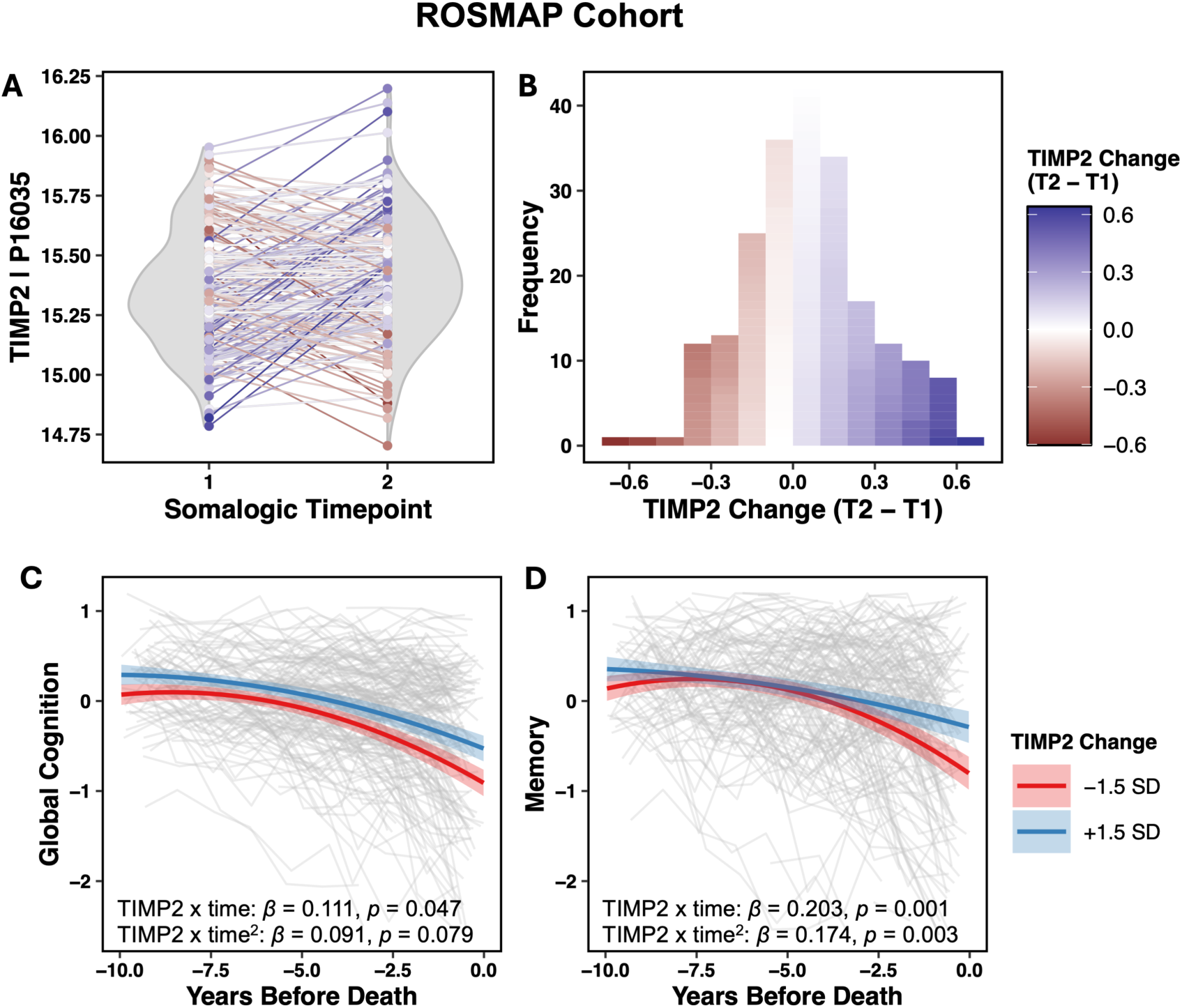
Changes in TIMP2 are associated with longitudinal cognitive trajectories. Longitudinal TIMP2 data in ROSMAP **(E)** show that change in TIMP2 (Timepoint 2 - Timepoint 1) is normally distributed **(F)** and that decreases in plasma TIMP2 are associated with steeper cognitive declines over time, particularly in memory **(G,H)**.

### Plasma TIMP2 Levels Are Associated with Enriched Lifestyle in Humans

We next tested whether plasma TIMP2 levels associated with established modifiable, neuroprotective behaviors. Using wearable actigraphy monitoring in the UCSF cohort, higher average daily step count associated with higher plasma TIMP2 (*Std. β*=0.239, *95%CI*=0.011 to 0.466, *p*=0.040; **Fig. 4A**; **Supplementary Table 5**). In the longitudinal ROSMAP cohort, changes in a self-reported multi-domain lifestyle composite (including physical activity, social activity, cognitive activity, and “life space” representing extent of movement through one’s environment (23)) were positively associated with change in TIMP2 levels over time (*Std. β*=0.193, *95%CI*=0.052 to 0.338, *p*=0.008; **Fig. 4B**; **Supplementary Table 6**; and see **Suppl. Fig. 2** for associations between TIMP2 and each self-reported lifestyle factor).

**Fig. 4.**
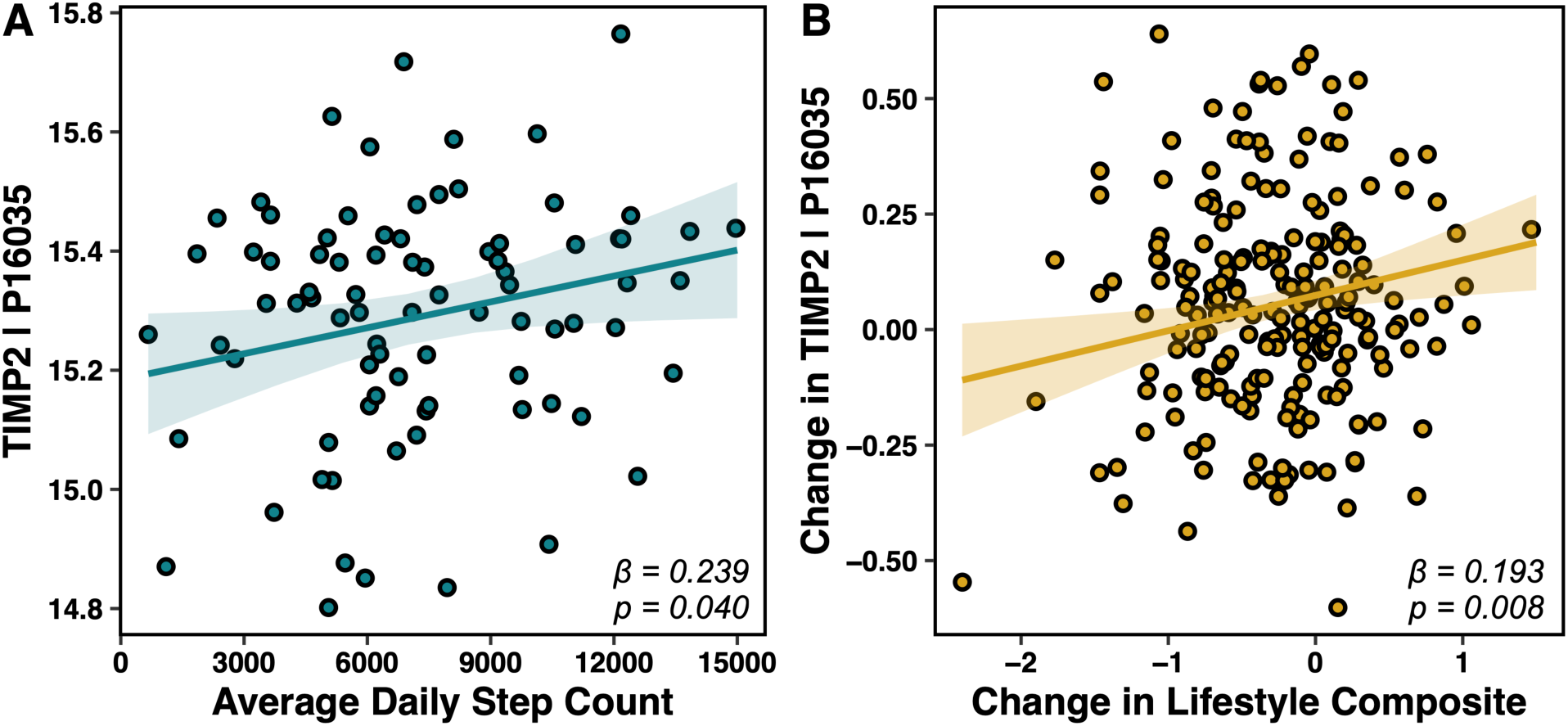
TIMP2 relates to neuroprotective lifestyle behaviors cross-sectionally and longitudinally. **(A)** Higher levels of physical activity associated with higher plasma TIMP2 concentrations in the UCSF cohort and **(B)** changes in engagement in multi-domain lifestyle factors associated with changes in plasma TIMP2 concentrations over time in the longitudinal ROSMAP cohort.

### Environmental enrichment increases plasma TIMP2 levels and adult hippocampal neurogenesis in WT but not TIMP2 KO mice

Given the association between plasma TIMP2 levels and social and environmental enrichment we observed in human subjects, we sought to test the potential for plasma TIMP2 modulation via behavior in an animal model. We adapted paradigms established to study the effects of environmental enrichment on synaptic plasticity in mice (27–29) to test whether enrichment exposure influences plasma TIMP2 levels. In this approach (**Fig. 1C**), mice are co-housed while exposed to various toys and a running wheel, the arrangement of which was regularly rotated each day to maintain novelty across the experiment. Wildtype (WT) mice aged for 2-3 months were exposed daily to 6 hours of the enriched environment (EE) or standard environment (SE) paradigm for 21 days, after which blood was collected for plasma TIMP2 measurements by immunoblotting (**Fig. 5A**). Female mice exposed to EE for 3 weeks exhibited significantly elevated plasma TIMP2 levels relative to those exposed to SE conditions (**Fig. 5B-C**), supporting a direct link between enrichment exposure and levels of blood-borne factors. We performed the same analysis in a separate cohort of male WT mice (**Suppl. Fig. 3A-B**), revealing a similar effect of enrichment on increasing plasma TIMP2 levels, though the result did not reach statistical significance (*P*=0.06), perhaps reflecting subtle sex-associated effects of enrichment on circulating TIMP2 levels.

**Fig. 5.**
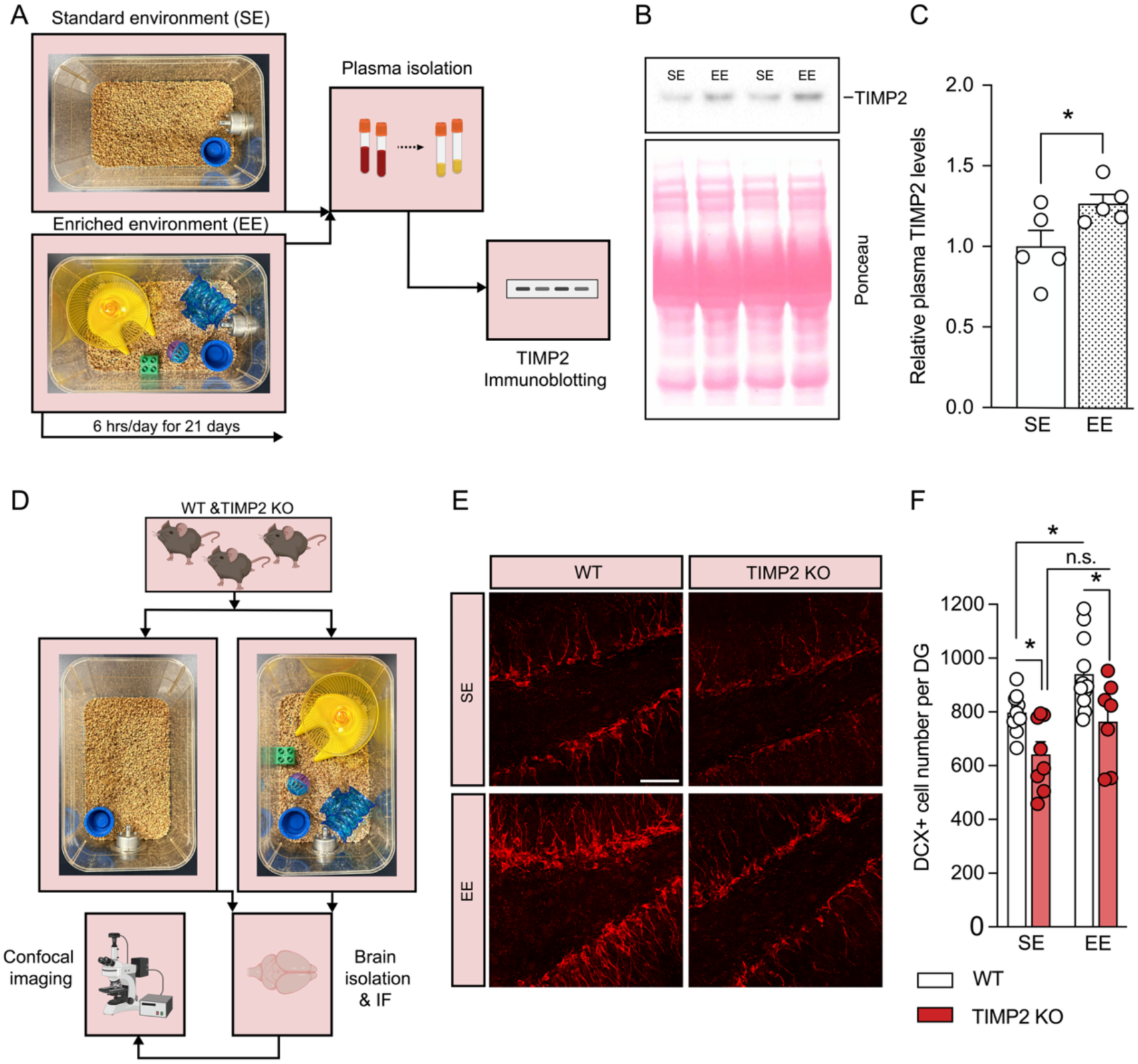
Enriched housing conditions increase plasma TIMP2 levels and promote hippocampal neurogenesis in a TIMP2-dependent manner. **(A)** Schematic diagram of the enriched environment (EE) or standard environment (SE) paradigm for wildtype (WT) mice using actual images of sample enrichment or standard configurations, after which blood is collected for plasma immunoblotting experiments. **(B)** TIMP2 immunoblotting (top) from plasma of WT mice exposed to EE and SE paradigm with Ponceau S stain (bottom) (N = 5 female mice per group; 2–3 months of age) with **(C)** corresponding quantification. \**P*<0.05, Student’s t test. **(D)** Schematic diagram of the EE and SE workflow used for WT and TIMP2 KO mice to examine adult hippocampal neurogenesis by confocal imaging. **(E)** Representative confocal images of brain sections containing dentate gyrus (DG) from WT and TIMP2 KO female mice that were stained with antibodies for doublecortin (DCX) exposed to SE or EE with **(F)** corresponding quantification of the total number of DCX⁺ cells in DG in each group (N = 7–11 mice per group, 2–3 months of age; 2-way ANOVA, Sidak’s test for multiple comparisons; \**P* < 0.05; n.s., not significant. Data represent mean ± SEM, with individual mice represented by points.

Our recent work described a role for TIMP2 in regulating adult neurogenesis within the subgranular zone of the (DG) gyrus (7), and our earlier work demonstrated a role for blood-borne TIMP2 in regulating hippocampal plasticity in aged contexts (6). Given these links, and the large body of evidence establishing that enrichment in environmental conditions improves hippocampal plasticity, especially adult neurogenesis (31), we sought to examine the relationship between TIMP2 and the effect of enrichment on neurogenesis using our enrichment paradigm. Specifically, we tested the hypothesis that the pro-neurogenic effects of EE on the adult hippocampus are dependent on the presence of TIMP2. We generated female WT and TIMP2 knockout (TIMP2 KO) littermates at 2-3 months of age, a timepoint at which plasma TIMP2 levels are high (6), and exposed the mice to the EE and SE paradigm described previously (**Fig. 1D**). Following the conclusion of these exposures (**Fig. 5D**), we isolated hemibrains and applied confocal imaging for quantification of the number of immature neuroblasts in brain sections containing hippocampus that were immunostained with doublecortin (DCX) antibody (**Fig. 5E**). As expected, EE exposure significantly increased the number of DCX⁺ cells in the DG compared to SE (**Fig. 5F**) in WT mice, demonstrating enrichment-driven enhancement of adult neurogenesis and validating the paradigm. We also verified the decline in adult neurogenesis in mice lacking TIMP2 relative to WT mice, as we previously reported (7) (**Fig. 5F**). Supporting our hypothesis, TIMP2 KO mice did not exhibit an elevation in DCX+ cell number following EE exposure (**Fig. 5F**), arguing that the pro-neurogenic response to enrichment is blocked by loss of TIMP2.

Our results demonstrate that EE increases circulating TIMP2 in mice and that TIMP2 is necessary for the positive effects of EE conditions on adult hippocampal neurogenesis. These findings support a link between higher levels of TIMP2 and its role as a molecular mediator of hippocampal plasticity via environmental enrichment experience.

## DISCUSSION

This translational study integrated findings from multiple independent human cohorts and a novel enrichment paradigm in mice to support TIMP2 as a human-relevant brain rejuvenation factor implicated in the link between modifiable lifestyle factors and brain health. By leveraging both cross-sectional and longitudinal data, we extend previous preclinical findings to humans, demonstrating that higher plasma TIMP2 levels are robustly associated with larger brain volumes and better cognitive performance. Stable-to-increasing plasma TIMP2 levels predicted more stable memory trajectories over time, supporting its use as a potential biomarker of cognitive resilience. Additionally, our findings are the first to demonstrate that TIMP2 is associated with multi-domain modifiable lifestyle behaviors in humans. Complementary mouse studies showed that circulating TIMP2 levels are elevated following exposure to an enriched environment, providing experimental support for behaviorally-driven regulation of this protein. Our results using TIMP2 knockout mice reveal that some plasticity benefits conferred by environmental enrichment are mediated by TIMP2. Together, these results position TIMP2 as a factor underlying neuroprotective effects of lifestyle behaviors that highlight its translational potential as a therapeutic target for healthy brain aging and dementia prevention.

These findings align with growing evidence implicating TIMP2 in immunovascular (32, 33) and ECM regulation (34, 35), processes disrupted in aging, injury, and neurodegeneration (36, 37). TIMP2 is primarily known for its regulation of its canonical binding partner, matrix metalloproteinase 2 (MMP2), though some MMP-independent roles have been demonstrated in the CNS (38). Recent work highlights its role in broader ECM and neuroinflammatory signaling pathways in the CNS, as it is highly expressed by neurons, decreases ECM accumulation in synapses to support neuroplasticity (6, 7), and reduces microglia activation (39). Genetic support for the relevance of TIMP2 in human brain aging is supported by a recent study showing positive associations between a polygenic proxy of circulating TIMP2 levels and cognitive functioning (40). Further, TIMP2 signaling has also been implicated in neurodegeneration (41), including altered TIMP2 expression alongside other extracellular matrix proteins in AD brain tissue and cerebrospinal fluid (42), though more work is needed to examine this association.

Notably, TIMP2 has also been linked to angiogenic and vascular remodeling pathways in humans relevant to exercise-associated brain benefits (43). Our group recently reported associations between increased physical activity and upregulation of peripheral proteins involved in vascular remodeling, cell adhesion, and ECM signaling, including TIMP2 (44). Our current mouse results further support the hypothesized directionality of these associations and that TIMP2 may be modulated through lifestyle intervention. Findings support future work examining other blood-borne factors linked to modifiable behaviors and brain health, including ANTXR2 and Gpld1 (44–46), as there are likely multiple underlying pathways involved. Overall, converging lines of evidence support a model in which TIMP2 acts as a behavior-responsive, neuroprotective circulating factor.

Several limitations warrant consideration. Sample sizes in our human cohorts were modest; however, replication of results across three independent cohorts strengthens generalizability and robustness of our findings. Additionally, orthogonal measurement of TIMP2 across assays in human plasma increases confidence of the biological target quantified in humans. Given recent identification of additional putative and non-canonical TIMP2 binding proteins (47), future work is needed to understand the impact of the TIMP2 interactome in the phenotypes observed and the extent to which these may also be modifiable through lifestyle interventions.

Together, these findings provide translational evidence for plasma TIMP2 as a behavior-responsive, neuroprotective factor relevant to human brain aging. By linking TIMP2 to both neural and cognitive markers of brain aging, as well as to neuroprotective lifestyle behaviors, our data support TIMP2 as a pathway through which lifestyle influences cognitive aging through the periphery. These findings also underscore the potential of TIMP2 as both a readout for monitoring the neuroprotective impact of lifestyle interventions and support its investigation as a direct therapeutic target for prevention of cognitive decline in older adults.

## Funding & Acknowledgements

We would like to thank clinical coordinators, staff, study subjects at UCSF, Stanford ADRC, and the Rush ADC, as well as S. Philippi (ISMMS) for assistance with figure illustration; components of some figure panels used BioRender content. This work was supported by the National Institute on Aging (K23AG084883 (EWP), K23AG090757 (RS), R01AG032289 (JHK), R01AG048234 (JHK), P30AG062422 (GDR), R01AG072475 (KBC), K23AG058752 (KBC), K23AG09073301A1 (KY), R01AG061382 (JMC), RF1AG072300 (JMC), P30AG10161 (DAB), P30AG72975 (DAB, R01AG17917 (DAB), and R01AG015819(DAB), the Larry Hillblom Foundation (2024-A-001-CTR; KBC), the Shenandoah Foundation (EWP), New Vision Research (CCAD 2024-001-1 (RS)), Alzheimer’s Association (AARF-23-1145318 (RS), AACSF-24-1307411 (KY)), the Knight Initiative for Brain Resilience (KY), Schwab Charitable (KY), the Iqbal and Asad Jamal Fund (KY), the Erb Family Foundation (KY), fellowships from American Academy of Neurology (RS), American Brain Foundation (RS), and Association for Frontotemporal Degeneration (RS). ROSMAP SomaScan data were also supported by Gates Ventures, Johnson & Johnson, and the Phil and Penny Knight Initiative for Brain Resilience.

## Data Availability

All data produced in the present study are available upon reasonable request to the UCSF ADRC, Stanford ADRC, or Rush ADC.

